# Germline Shelterin Loss of Function Predisposes to Lipoma and Uterine Leiomyoma: Population Evidence Bearing on Shelterin-Associated Sarcoma Risk

**DOI:** 10.64898/2026.11.03.26367209

**Authors:** Benjamin D. Lin, Karly Hoi-Lam Chan, Zheyang Zhang, Lillian Zhi, Andreana Leow, Joyce Camellia Chi, Maggie Zhou, Nam Bui, Kristen Ganjoo, Minggui Pan

**Author notes:** Corresponding author: Minggui Pan, MD, PhD.

## Abstract

**Purpose:** Germline shelterin variants were linked to sarcoma in a case-enriched cohort. Whether shelterin loss of function predisposes to mesenchymal neoplasia in the general population is unknown.

**Methods:** We analyzed rare-variant collapsing associations for five shelterin genes across neoplasm phenotypes in UK Biobank exomes, with Genebass as a second pipeline. Prespecified shelterin and centrosome gene sets were tested by permutation-calibrated statistics against a *TP53*/*RB1*/*NF1* positive control. Carrier risks were compared with telomere-lengthening predictions.

**Results:** Lipoma risk was increased in *TERF1* (odds ratio [OR], 3.32; *P* = 8.4 × 10⁻⁸), *POT1*, and *TERF2IP* carriers, and uterine leiomyoma risk in all four core shelterin genes, most strongly *POT1* (OR, 2.23; *P* = 4.0 × 10⁻⁵); POT1 and TINF2 reproduced in Genebass. *TERF1* was also associated with thyroid cancer and monoclonal gammopathy. Neither gene set nor the positive control was associated with self-reported sarcoma. Telomere lengthening accounted for *POT1*-associated chronic lymphocytic leukemia risk but not myeloma or *TINF2*-associated thyroid cancer, where observed risk exceeded prediction five- to seven-fold.

**Conclusion:** Shelterin loss of function is a population-level risk factor for lipoma and uterine leiomyoma, the benign counterparts of liposarcoma and leiomyosarcoma. *TERF1* joins *POT1* and *TINF2* as long-telomere cancer genes. Sarcoma-specific penetrance remains untested.

## Introduction

Sarcomas have the highest heritable fraction of any adult solid tumor, yet most familial and early-onset cases remain unexplained after testing for *TP53* and the classic predisposition syndromes.^1^ Whole-genome sequencing of the International Sarcoma Kindred Study recently implicated rare variants in the shelterin complex (*POT1*, *TINF2*, *TERF1*, *TERF2IP*) and in centrosome genes (*CEP63*, *CEP72*, *HAUS4*, *HAUS5*).^2^ These genes now appear on expanded sarcoma panels, but the association derives from a case-enriched cohort and has not been replicated.

Shelterin caps chromosome ends and restrains telomerase; its loss of function lengthens telomeres.^3–5^ Telomere-lengthening *POT1* and *TINF2* variants cause familial melanoma, chronic lymphocytic leukemia (CLL), thyroid cancer, glioma, and cardiac angiosarcoma.^6–12^ Population studies have since established three points: rare *POT1*, *TERF1*, and *TINF2* truncating variants lengthen leukocyte telomere length (LTL) in UK Biobank;^13^ *POT1* carriers in UK Biobank have an eight-fold risk of lymphoid malignancy;^12^ and genetically longer telomeres cause CLL, melanoma, thyroid cancer, and myeloma.^14^ Uterine fibroids have been noted in *POT1* long-telomere kindreds.^12^

Three questions remain open. First, whether the shelterin spectrum extends to mesenchymal neoplasia, which would give the sarcoma association a lineage-specific basis. Second, whether *TERF1* and *TERF2IP*, which lengthen telomeres but lack established cancer phenotypes, share the spectrum. Third, whether telomere length alone accounts for carrier risk. We addressed these questions, and tested the proposed sarcoma genes directly, using population exome and genome-wide association resources.

## Materials and Methods

### Data Sources

This study used publicly available summary statistics. Gene-level rare-variant associations came from UK Biobank whole-exome sequencing, analyzed through the AstraZeneca PheWAS portal^15^ and, as a second analytic pipeline, Genebass.^16^ Outcome genome-wide association statistics came from FinnGen release 12.^17^ Institutional review was not required for deidentified summary data. The two resources analyze substantially the same individuals: 459,449 participants of European ancestry in the AstraZeneca release used here and 394,841 in Genebass, drawn from the same UK Biobank exome tranche, so the Genebass sample is largely contained within the AstraZeneca sample. Agreement between them therefore tests variant masks, quality control, and test statistics rather than sampling.

### Phenome-Wide Collapsing Analysis

We extracted results for *POT1*, *TERF1*, *TERF2*, *TERF2IP*, and *TINF2* across all neoplasm-related binary phenotypes (ICD-10 cancer registry, hospital inpatient, death registry, self-report, histology, and union definitions) and LTL in individuals of European ancestry. The portal export retained gene–model– phenotype results at *P* ≤ .10 (1,118 phenotypes). Four collapsing models were considered: protein-truncating variants (PTV); PTV plus rare damaging missense (PTV + RDM); ultra-rare variants; and synonymous variants as a negative control. Binary associations are ORs with 95% CIs (Fisher’s exact test). The prespecified mesenchymal outcomes were benign lipomatous neoplasm (ICD-10 D17), uterine leiomyoma (D25), soft-tissue malignancy (C49), mesothelial and soft-tissue malignancy (C45-C49), bone malignancy (C40-C41), and connective tissue histology (ICD-O-3 8800-9370). Union definitions (any source) were primary. We treat *P* < 1 × 10⁻⁵ as robust in this five-gene analysis, *P* < .05 as nominal, and concordance across genes as supporting evidence.

### Second Pipeline

For uterine leiomyoma (D25; 9,560 cases) and 37 ICD-10 cancer sites, we retrieved Genebass pLoF and synonymous burden and SKAT-O^18^ results for the six shelterin genes (adding *ACD*).

### Gene-Set Tests of the Proposed Sarcoma Genes

From Genebass, we retrieved exome-wide gene-level results for self-reported sarcoma or fibrosarcoma (208 cases), thyroid cancer (C73; 340 cases), and melanoma (C43; 2,024 cases) for pLoF, pLoF plus missense, and synonymous masks. Seven sets were prespecified: shelterin core (*POT1*, *TERF1*, *TINF2*, *TERF2IP*), all six shelterin genes, an extended telomere set (adding *SMARCAL1*, *TIMELESS*, *STAG3*, *CTC1*, *NHP2*), centrosome core (*CEP63*, *CEP72*, *HAUS4*, *HAUS5*), an extended centrosome set (adding *CEP89*, *SSNA1*, *PCM1*, *CEP57*, *MZT1*), the combined core, and a positive control (*TP53*, *RB1*, *NF1*), mirroring the discovery study.^2^ Sets were tested with a directional Stouffer statistic of signed burden *z* scores and a nondirectional aggregated Cauchy association test (ACAT)^19^ of SKAT-O *P* values.

### Observed Versus Telomere-Predicted Risk

We estimated the causal effect of LTL on CLL, thyroid cancer, melanoma, and multiple myeloma by two-sample Mendelian randomization (MR), using 101 LTL instruments (mean F, 134)^20^ and FinnGen R12 outcomes. The primary estimator was inverse-variance weighted (IVW) MR,^21^ with MR-Egger,^22^ weighted median,^23^ and exclusion of telomere-biology loci as sensitivity analyses. For each lead gene– cancer pair, we computed the risk predicted by telomere lengthening alone as exp(ΔLTL × βIVW), where ΔLTL is the carrier LTL difference in SD units and βIVW the log OR per SD, with 95% CIs from the MR standard error. Observed-to-predicted ratios above 1 indicate risk not explained by telomere length under a log-linear model.

### Statistical Analysis

Analyses used Python 3 with pandas, NumPy, SciPy, and matplotlib; the permutation seed was 2026. *P* values are two-sided except the directional gene-set statistic, which is one-sided for risk; ACAT is nondirectional. All analyses are complete-case on published summary statistics.

The prespecified mesenchymal family comprised six phenotypes, five genes, and four collapsing models (120 tests; Bonferroni α = 4.2 × 10⁻⁴). Because the models are nested and union phenotypes contain their source-specific components, these tests are correlated and Bonferroni is conservative. We therefore prespecified *P* < 1 × 10⁻⁵ as robust, *P* < 4.2 × 10⁻⁴ as family-wise significant, and *P* < .05 as nominal, and required concordance across genes or across pipelines before interpreting a nominal result. Because the portal export truncated at *P* ≤ .10, genomic inflation factors and false-discovery rates could not be computed, and results absent from the export are reported as unavailable rather than as null.

Synonymous collapsing models were a within-gene negative control, and *TP53*, *RB1*, and *NF1* a between-gene positive control for sarcoma. The AstraZeneca PheWAS and Genebass analyses draw on overlapping UK Biobank participants, so their agreement tests analytic robustness across different variant masks, quality control, and test statistics rather than replication in an independent sample.

For each phenotype and mask, gene-level burden estimates were converted to signed *z* scores, *z* = sign(β) × Φ⁻¹(1 − *P*/2), and combined as Σ*z*/√*k*. The null distribution was 100,000 random sets of *k* genes drawn without replacement from the 17,522 to 19,020 genes with non-missing burden results in that stratum, with empirical *P* = (*b* + 1)/(*B* + 1). Random sets were matched on gene count only, not on cumulative allele frequency or variant count. ACAT combined SKAT-O *P* values over 20,000 random sets. Genebass returns a SKAT-O *P* of exactly 1 for genes with too few qualifying variants (64.7% of genes for sarcoma, 52.0% for thyroid cancer, and 18.1% for melanoma), and one such gene saturates the Cauchy statistic; gene sets containing one are reported as not evaluable. The null is affected in the same way: 95% to 99.8% of random sets for thyroid cancer contain a saturated gene and sit at the ceiling, so an evaluable observed set appears extreme against them. ACAT was therefore calibrated against random sets drawn from evaluable genes only. Calibration against the whole gene pool, the conventional choice, returns *P* values two- to 549-fold smaller here and is anticonservative.

For Mendelian randomization, instrument strength was summarized by the mean *F* statistic, between-instrument heterogeneity by Cochran *Q*, and directional pleiotropy by the MR-Egger intercept; I²GX was 0.98, supporting the no-measurement-error assumption for MR-Egger. Sensitivity analyses excluded eight telomere-biology loci (93 instruments) and palindromic variants (86 instruments). Predicted ORs treat each gene’s ΔLTL as fixed and propagate only the MR standard error, so their CIs are narrower than the true uncertainty, and the log-linear extrapolation to carrier-sized LTL shifts is untested. Analysis code and the summary-statistic extracts analyzed are available as described under Data and Code Availability.

## Results

### Shelterin Loss of Function Predisposes to Lipoma and Uterine Leiomyoma

Benign adipocytic and smooth muscle tumors were among the strongest shelterin associations in the neoplasm phenome (Fig 1; Table 1). For lipoma (10,042 cases), *TERF1* PTV + RDM carriers had an OR of 3.32 (95% CI, 2.27 to 4.88; 30 carriers; *P* = 8.4 × 10⁻⁸), and *TERF1* PTV carriers an OR of 4.45 (95% CI, 2.40 to 8.25; *P* = 5.4 × 10⁻⁵). The *TERF1* signal was present in hospital-coded lipoma (OR, 3.63; *P* = 2.5 × 10⁻⁷) and was strongest for truncal subcutaneous (PTV OR, 8.74; *P* = 2.8 × 10⁻⁵) and intra-abdominal lipomas (PTV + RDM OR, 10.7; *P* = 3.0 × 10⁻⁵). *POT1* (PTV OR, 2.94; 95% CI, 1.75 to 4.95; *P* = 2.8 × 10⁻⁴) and *TERF2IP* (PTV OR, 2.40; 95% CI, 1.39 to 4.17; *P* = .006) carriers showed the same association; *TINF2* was associated only with truncal lipoma (PTV OR, 3.85; *P* = .047).

**FIG 1.**
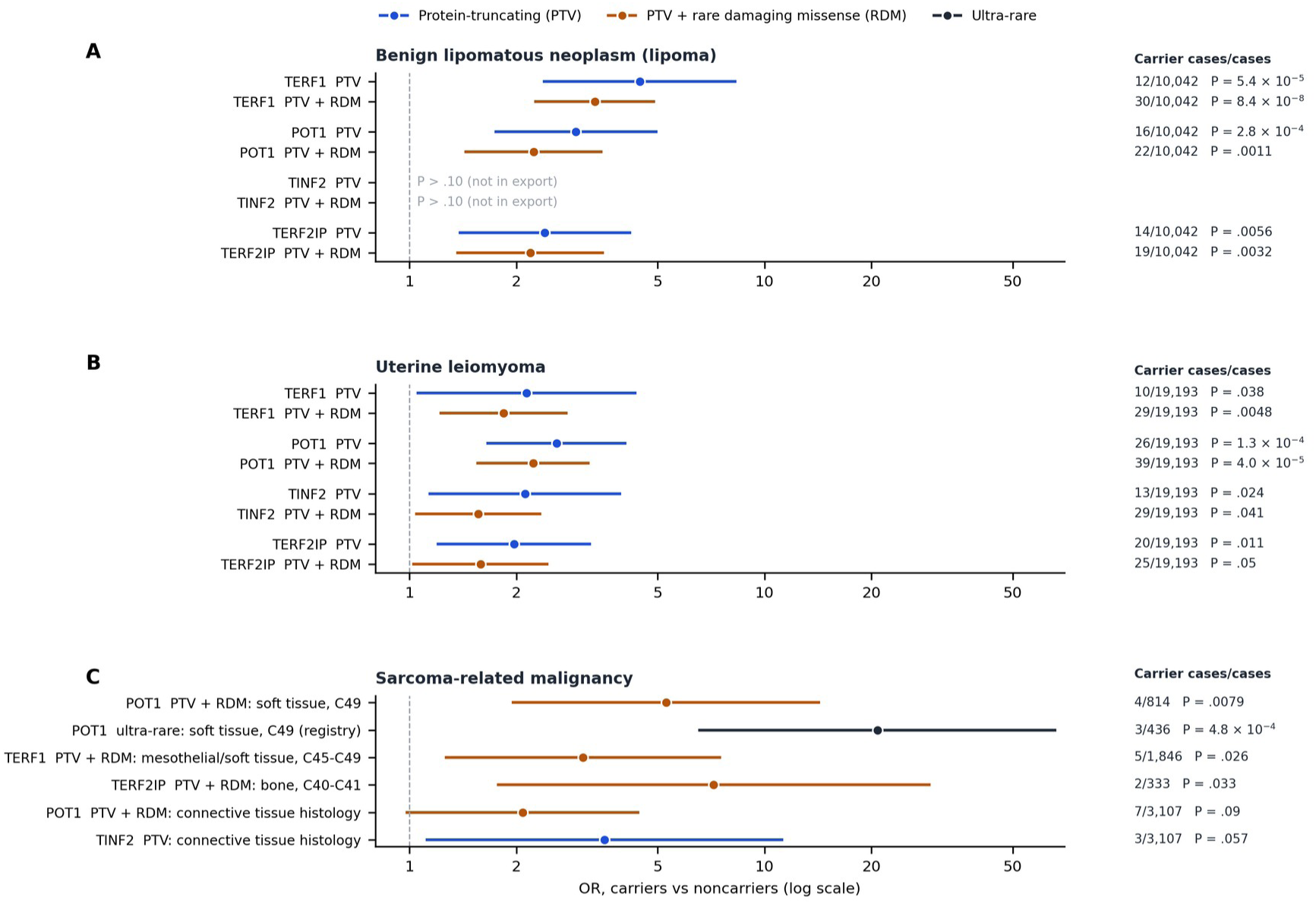
Shelterin loss of function and mesenchymal neoplasia in UK Biobank. ORs (log scale) with 95% CIs for carriers of protein-truncating variants (PTV; blue), PTV plus rare damaging missense variants (RDM; orange), and ultra-rare variants (black) versus noncarriers, European ancestry. (A) Benign lipomatous neoplasm (ICD-10 D17, union of sources; 10,042 cases). (B) Uterine leiomyoma (D25, union; 19,193 cases). (C) Sarcoma-related malignancy. Carrier cases/total cases and Fisher’s exact *P* values at right. Results with *P* > .10 were not available from the portal export.

**Table 1.** Mesenchymal and *TERF1* Neoplasm Associations of Rare Shelterin Variants in UK Biobank (European ancestry)

| Gene | Model | Phenotype (source) | Carrier cases/cases | OR (95% CI) | *P* | ΔLTL, SD (95% CI) |
|---|---|---|---|---|---|---|
| **Lipoma** |  |  |  |  |  |  |
| *TERF1* | PTV + RDM | Benign lipomatous neoplasm, D17 (union) | 30/10,042 | 3.32 (2.27 to 4.88) | 8.4 × 10<sup>−8</sup> | 1.07 (0.98 to 1.16) |
| *TERF1* | PTV | Benign lipomatous neoplasm, D17 (union) | 12/10,042 | 4.45 (2.40 to 8.25) | 5.4 × 10<sup>−5</sup> | 1.39 (1.23 to 1.54) |
| *TERF1* | PTV | Lipoma, skin and subcutis of trunk, D17.1 (union) | 7/2,968 | 8.74 (4.02 to 19.0) | 2.8 × 10<sup>−5</sup> | 1.39 (1.23 to 1.54) |
| *TERF1* | PTV + RDM | Lipoma, intra-abdominal organs, D17.5 (union) | 6/631 | 10.7 (4.73 to 24.1) | 3.0 × 10<sup>−5</sup> | 1.07 (0.98 to 1.16) |
| *POT1* | PTV | Benign lipomatous neoplasm, D17 (union) | 16/10,042 | 2.94 (1.75 to 4.95) | 2.8 × 10<sup>−4</sup> | 1.27 (1.15 to 1.39) |
| *TERF2IP* | PTV | Benign lipomatous neoplasm, D17 (union) | 14/10,042 | 2.40 (1.39 to 4.17) | .0056 | 0.22 (0.10 to 0.33) |
| *TINF2* | PTV | Lipoma, skin and subcutis of trunk, D17.1 (union) | 3/2,968 | 3.85 (1.21 to 12.2) | .047 | 0.68 (0.51 to 0.85) |
| **Uterine leiomyoma** |  |  |  |  |  |  |
| *POT1* | PTV + RDM | Uterine leiomyoma, D25 (union) | 39/19,193 | 2.23 (1.56 to 3.18) | 4.0 × 10<sup>−5</sup> | 0.94 (0.86 to 1.03) |
| *POT1* | PTV | Uterine leiomyoma, D25 (union) | 26/19,193 | 2.59 (1.66 to 4.05) | 1.3 × 10<sup>−4</sup> | 1.27 (1.15 to 1.39) |
| *POT1* | PTV | Leiomyoma, unspecified, D25.9 (hospital) | 16/6,339 | 4.62 (2.72 to 7.86) | 1.9 × 10<sup>−6</sup> | 1.27 (1.15 to 1.39) |
| *TERF1* | PTV + RDM | Uterine leiomyoma, D25 (union) | 29/19,193 | 1.84 (1.22 to 2.76) | .0048 | 1.07 (0.98 to 1.16) |
| *TERF2IP* | PTV | Uterine leiomyoma, D25 (union) | 20/19,193 | 1.97 (1.20 to 3.22) | .011 | 0.22 (0.10 to 0.33) |
| *TINF2* | PTV | Uterine leiomyoma, D25 (union) | 13/19,193 | 2.11 (1.14 to 3.91) | .024 | 0.68 (0.51 to 0.85) |
| **Sarcoma-related malignancy** |  |  |  |  |  |  |
| *POT1* | PTV + RDM | Soft-tissue malignancy, C49 (union) | 4/814 | 5.28 (1.96 to 14.2) | .0079 | 0.94 (0.86 to 1.03) |
| *POT1* | Ultra-rare | Soft-tissue malignancy, C49 (registry) | 3/436 | 20.8 (6.56 to 65.9) | 4.8 × 10<sup>−4</sup> | — |
| *TERF1* | PTV + RDM | Mesothelial and soft tissue, C45-C49 (union) | 5/1,846 | 3.08 (1.27 to 7.48) | .026 | 1.07 (0.98 to 1.16) |
| *TERF2IP* | PTV + RDM | Bone and articular cartilage, C40-C41 (union) | 2/333 | 7.19 (1.78 to 29.1) | .033 | 0.16 (0.06 to 0.25) |
| **Other *TERF1* associations** |  |  |  |  |  |  |
| *TERF1* | PTV + RDM | Thyroid cancer, C73 (union) | 6/1,045 | 6.91 (3.05 to 15.6) | 3.2 × 10<sup>−4</sup> | 1.07 (0.98 to 1.16) |
| *TERF1* | PTV + RDM | Benign thyroid neoplasm, D34 (hospital) | 4/398 | 11.5 (4.27 to 31.2) | 4.9 × 10<sup>−4</sup> | 1.07 (0.98 to 1.16) |
| *TERF1* | PTV | MGUS, D47.2 (hospital) | 3/297 | 34.9 (10.9 to 111) | 1.1 × 10<sup>−4</sup> | 1.39 (1.23 to 1.54) |
| *TERF1* | PTV | Lymphoid/ hematopoietic malignancy, C81-C96 (union) | 10/8,541 | 4.31 (2.21 to 8.41) | 2.6 × 10<sup>−4</sup> | 1.39 (1.23 to 1.54) |
| *TERF1* | PTV + RDM | Malignant neoplasm of choroid, C69.3 (hospital) | 3/182 | 18.0 (5.71 to 56.6) | 7.2 × 10<sup>−4</sup> | 1.07 (0.98 to 1.16) |

Abbreviations: MGUS, monoclonal gammopathy of undetermined significance; OR, odds ratio; PTV, protein-truncating variant; RDM, rare damaging missense. Source: AstraZeneca PheWAS portal; Fisher's exact test. ΔLTL, carrier-minus-noncarrier leukocyte telomere length in SD for the variant class shown in that row; it is a property of the gene and variant class rather than of the phenotype and therefore repeats across rows. Ultra-rare variants were not analyzed for telomere length.

For uterine leiomyoma (19,193 cases), all four core shelterin genes were associated in the risk direction: *POT1* (PTV + RDM OR, 2.23; 95% CI, 1.56 to 3.18; *P* = 4.0 × 10⁻⁵; PTV OR, 2.59; *P* = 1.3 × 10⁻⁴), *TINF2* (PTV OR, 2.11; *P* = .024), *TERF2IP* (PTV OR, 1.97; *P* = .011), and *TERF1* (PTV + RDM OR, 1.84; *P* = .005). In hospital-coded unspecified leiomyoma, the *POT1* PTV association reached *P* = 1.9 × 10⁻⁶ (OR, 4.62). Genebass analysis of leiomyoma (9,560 cases) reproduced the *POT1* (burden *P* = 5.1 × 10⁻⁵) and *TINF2* (SKAT-O *P* = 2.2 × 10⁻⁴) signals, with *TERF2IP* at *P* = .052 and *TERF1* at *P* = .12, whereas *TERF2* and *ACD*, which have little effect on telomere length, were null (*P* > .5). Synonymous variants were not associated with leiomyoma in either pipeline (Genebass *P* ≥ .30 for five of six genes).

Of eight gene–tumor pairs (four genes, two tumors), seven showed nominally significant PTV associations in the risk direction and the eighth was significant for a lipoma subsite. Risk did not scale with telomere lengthening: *TERF2IP* variants, which lengthen LTL by only 0.22 SD compared with 1.27 SD for *POT1* (Supplementary Figure S1), conferred similar leiomyoma and lipoma risks. Synonymous *POT1* variants showed weak, nonsignificant elevations for both tumors (OR, 1.46 and 1.38; *P* = .09 and .08), which we note as a caution for this gene.

### TERF1 Is a Long-Telomere Cancer Gene

*TERF1* produced the largest telomere elongation of any shelterin gene (PTV, +1.39 SD; 95% CI, 1.23 to 1.54; *P* = 2.2 × 10⁻⁶⁹) and a cancer spectrum overlapping that of *POT1* and *TINF2* (Table 1). *TERF1* PTV + RDM carriers had increased risk of thyroid cancer (OR, 6.91; 95% CI, 3.05 to 15.6; *P* = 3.2 × 10⁻⁴), with concordant self-reported thyroid cancer (OR, 9.66; *P* = 9.3 × 10⁻⁴) and benign thyroid neoplasm (OR, 11.5; *P* = 4.9 × 10⁻⁴). *TERF1* PTV carriers had increased risk of monoclonal gammopathy of undetermined significance (MGUS; OR, 34.9; 95% CI, 10.9 to 111; 3 of 297 cases; *P* = 1.1 × 10⁻⁴) and lymphoid and hematopoietic malignancy (OR, 4.31; *P* = 2.6 × 10⁻⁴). *TERF1* PTV + RDM carriers also had increased risk of choroidal malignancy (OR, 18.0; 3 of 182 cases; *P* = 7.2 × 10⁻⁴), a site dominated by uveal melanoma. Across all benign neoplasms, *TERF1* PTV + RDM carriers had an OR of 1.86 (*P* = 2.3 × 10⁻⁸). The previously described *POT1* and *TINF2* associations with CLL, myeloma, thyroid cancer, and melanoma were reproduced (Supplementary Figure S2), and shelterin pLoF variants were nominally associated with 12.6% of 222 gene–site tests across 37 cancer sites, compared with 4.5% for synonymous variants (OR, 3.06; *P* = .003).

### The Proposed Sarcoma Genes Could Not Be Tested at Population Scale

For malignant mesenchymal outcomes, shelterin associations were consistent in direction with the benign tumors but imprecise (Fig 1C). *POT1* PTV + RDM carriers had an OR of 5.28 for soft-tissue malignancy (95% CI, 1.96 to 14.2; 4 of 814 cases; *P* = .008), and ultra-rare *POT1* variants an OR of 20.8 (3 of 436 registry cases; *P* = 4.8 × 10⁻⁴). *TERF1* was associated with mesothelial and soft-tissue malignancy (OR, 3.08; 5 of 1,846; *P* = .026) and *TERF2IP* with bone malignancy (OR, 7.19; 2 of 333; *P* = .033). For histology-coded connective tissue malignancy (3,107 cases), *POT1* (OR, 2.08; 95% CI, 0.98 to 4.40) and *TINF2* (OR, 3.54; 95% CI, 1.12 to 11.2) estimates were elevated but did not reach significance.

Prespecified gene-set tests against self-reported sarcoma (208 cases) were null for every set (Table 2): shelterin core *z* = −0.59 (empirical *P* = .84) and centrosome core *z* = −0.01 (*P* = .29). The positive-control set of *TP53*, *RB1*, and *NF1* was equally null (*z* = −0.25; *P* = .39), establishing that this outcome cannot detect established sarcoma genes. Centrosome sets also lacked a directional pLoF signal for thyroid cancer and melanoma; the only nominal directional result, for the extended centrosome set with missense variants in thyroid cancer (*z* = 2.77; *P* = .003), was unaccompanied by a pLoF signal and would not survive correction for 63 tests. Nondirectional ACAT tests were no stronger: against a matched null, shelterin-containing sets reached empirical *P* values of .15 to .33 for thyroid cancer and .034 to .083 for melanoma.

**Table 2.**
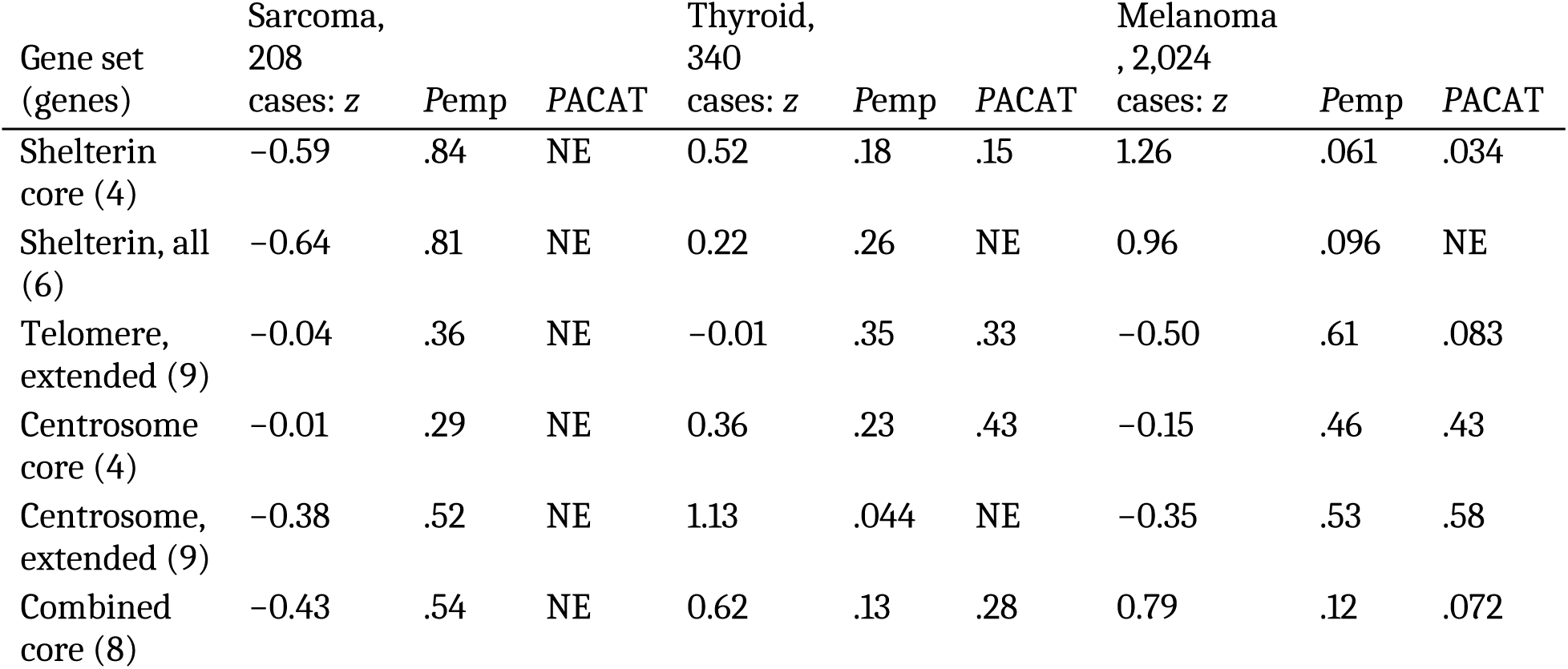
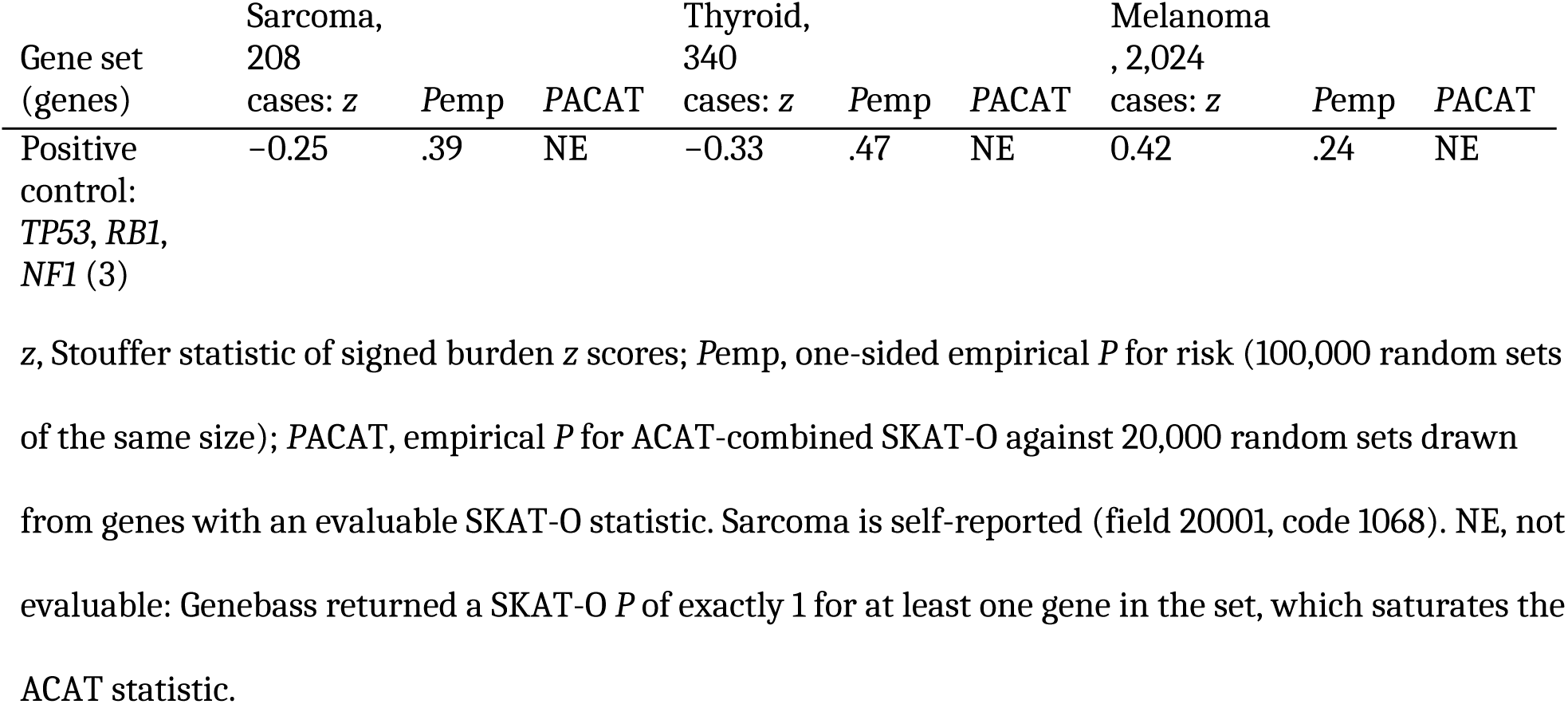
Gene-Set Tests of Predicted Loss-of-Function Burden for the Proposed Sarcoma Genes.

### Telomere Length Explains Some but Not All Carrier Risk

MR confirmed that longer LTL increases risk of CLL (IVW OR per SD, 4.35; 95% CI, 2.95 to 6.42), thyroid cancer (2.95), melanoma (2.28), and myeloma (1.79). Estimates were directionally consistent across MR-Egger and weighted-median estimators, although the myeloma effect was attenuated when telomere-biology loci were excluded (OR, 1.44; 95% CI, 0.98 to 2.10) and Cochran Q indicated heterogeneity for all outcomes except CLL (Supplementary Figure S3; Table 3). Applying these estimates to each gene’s LTL shift, telomere lengthening fully accounted for *POT1*-associated CLL (observed OR, 7.42; predicted, 6.46; ratio, 1.15) and was compatible with the *TINF2*–melanoma and *TERF1*–thyroid associations (ratios, 1.7 and 2.2; both ratio intervals include 1) (Fig 2). In contrast, observed risk exceeded telomere-predicted risk for *POT1*-associated myeloma (11.5 *v* 2.09; ratio, 5.5; 95% CI, 2.3 to 13.1) and *TINF2*-associated thyroid cancer (14.5 *v* 2.09; ratio, 7.0; 95% CI, 2.2 to 22.5), both ratio intervals excluding 1. The similar leiomyoma and lipoma risks across genes with a six-fold range in telomere elongation point in the same direction.

**FIG 2.**
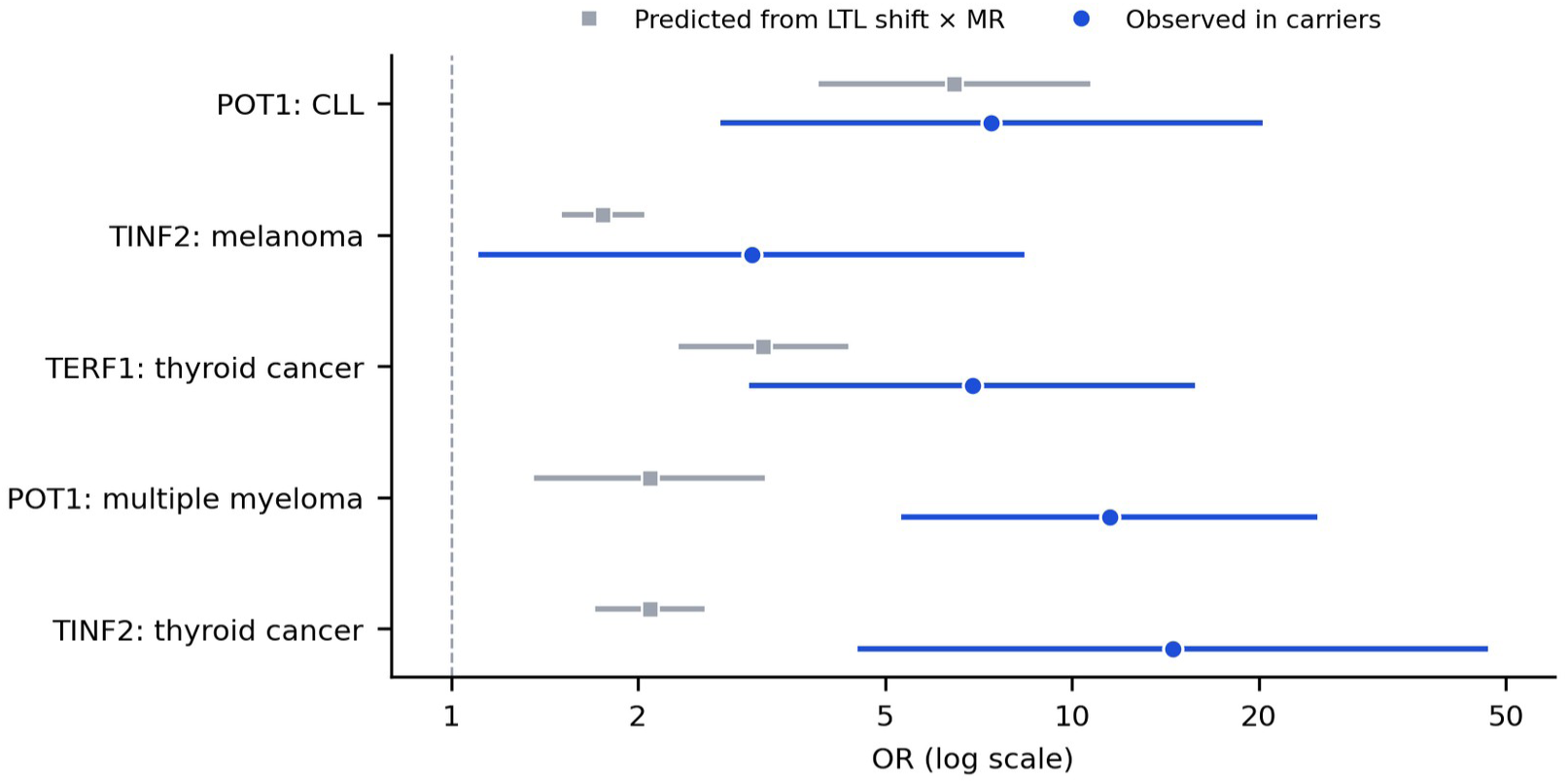
Observed carrier risk versus risk predicted from telomere lengthening. For each lead gene–cancer association, gray squares show the OR predicted from the carrier leukocyte telomere length shift multiplied by the Mendelian randomization estimate per SD (FinnGen R12), and blue circles show the observed carrier OR in UK Biobank. Bars, 95% CIs; log scale.

**Table 3.** Observed Carrier Risk Versus Risk Predicted From Telomere Lengthening.

| Gene: cancer | Carrier cases/cases | Carrier ΔLTL, SD | MR OR per SD (95% CI) | Predicted OR (95% CI) | Observed OR (95% CI) | Observed/ predicted (95% CI) |
|---|---|---|---|---|---|---|
| POT1: CLL | 4/949 | 1.27 | 4.35 (2.95 to 6.42) | 6.46 (3.95 to 10.6) | 7.42 (2.74 to 20.1) | 1.15 (0.38 to 3.49) |
| TINF2: melanoma | 4/5,041 | 0.68 | 2.28 (1.85 to 2.81) | 1.76 (1.52 to 2.02) | 3.05 (1.12 to 8.30) | 1.74 (0.63 to 4.78) |
| TERF1: thyroid cancer | 6/1,045 | 1.07 | 2.95 (2.22 to 3.92) | 3.18 (2.35 to 4.31) | 6.91 (3.05 to 15.6) | 2.17 (0.91 to 5.19) |
| POT1: multiple myeloma | 7/1,150 | 1.27 | 1.79 (1.28 to 2.48) | 2.09 (1.37 to 3.17) | 11.5 (5.37 to 24.6) | 5.51 (2.31 to 13.13) |
| TINF2: thyroid cancer | 3/774 | 0.68 | 2.95 (2.22 to 3.92) | 2.09 (1.72 to 2.54) | 14.5 (4.56 to 46.3) | 6.95 (2.15 to 22.50) |

ΔLTL, carrier minus noncarrier leukocyte telomere length for the variant class used (PTV, except *TERF1* PTV + RDM). MR, inverse-variance weighted Mendelian randomization, FinnGen R12 (101 instruments). Predicted OR = exp(ΔLTL × βMR). The interval on the ratio combines the observed and predicted standard errors on the log scale, treating the two estimates as independent and ΔLTL as fixed, so it is narrower than the true uncertainty. Phenotype sources: CLL, C91.1 (registry); melanoma, C43 (registry); TERF1–thyroid cancer, C73 (union); myeloma, C90 (registry); TINF2–thyroid cancer, C73 (registry). Full MR sensitivity analyses are in Supplementary Table S2 and Fig S3.

## Discussion

In roughly 450,000 exomes, shelterin loss of function predisposed to lipoma and uterine leiomyoma, the benign counterparts of liposarcoma and leiomyosarcoma. The association was shared by all four core shelterin genes, reproduced for leiomyoma in a second analytic pipeline, and absent for synonymous variants and for the shelterin genes with minimal telomere effects. *TERF1*, the gene with the largest telomere effect, also conferred risk of thyroid cancer, MGUS, and lymphoid malignancy, placing it beside *POT1* and *TINF2* as long-telomere cancer genes. The sarcoma and centrosome associations reported from a case-enriched cohort could not be tested adequately, because the sarcoma outcome lacked power even for positive-control genes.

These findings give the reported shelterin–sarcoma association a lineage-specific basis.^2^ Leiomyosarcoma and dedifferentiated liposarcoma are the most common adult soft-tissue sarcomas, and both depend on telomere maintenance through *TERT* activation or *ATRX*/*DAXX*-associated alternative lengthening of telomeres. A germline state that expands smooth muscle and adipocytic precursor pools, as the leiomyoma and lipoma associations imply, would enlarge the substrate for malignant transformation in precisely these lineages. The direction and magnitude of the malignant soft-tissue estimates (ORs, 3 to 5 for *POT1* and *TERF1*) are consistent with this model, although each rests on four or five carriers. The model predicts that shelterin carriers with sarcoma will be enriched for leiomyosarcoma and liposarcoma histology, and that their tumors will less often require somatic *TERT* or ALT activation; both predictions are testable in sequenced sarcoma cohorts.

The mesenchymal associations extend a recognized clinical observation. Uterine fibroids were noted in *POT1* long-telomere kindreds,^12^ and our data show that this is a population-level phenotype shared by all four core shelterin genes, with an approximately two-fold risk in carriers of each; TERF2 and ACD, which barely alter telomere length, were null. Lipoma has not, to our knowledge, been linked to germline telomere biology. The strength of the *TERF1*–lipoma association (*P* = 8.4 × 10⁻⁸), its presence in hospital-coded records, and its concentration in truncal and intra-abdominal sites argue against ascertainment artifact. Intra-abdominal lipomatous tumors deserve attention because well-differentiated liposarcoma can be coded as lipoma at that site.

Our leiomyoma results have to be read against a prior exome-wide search of the same resource. Välimäki et al. tested approximately 19,000 genes against the identical AstraZeneca portal endpoint under ten collapsing models and reported eight genes meeting odds ratio > 2.0 with *P* < 1 × 10⁻⁴, all converging on the SRCAP chromatin-remodeling complex; no shelterin gene was among them.^24^ The difference is case accrual, not effect size. Their release held 15,780 cases of this endpoint against the 19,193 analyzed here. Rescaling our *POT1* carrier counts to their case and control totals leaves the odds ratio unchanged (2.22 v 2.23) but moves *P* to 1.1 × 10⁻⁴ for PTV + RDM and 3.8 × 10⁻⁴ for PTV, both outside their reporting threshold. Shelterin and SRCAP are therefore best read as independent moderate-penetrance routes to uterine leiomyoma, the shelterin route distinguished by its extension to extrauterine adipocytic neoplasia.

Telomere length did not account for all carrier risk. The *POT1*–CLL association matched the risk predicted from telomere lengthening, consistent with prolonged B-cell replicative capacity driving clonal expansion.^5,12,25^ But *POT1*-associated myeloma and *TINF2*-associated thyroid cancer^26^ exceeded predictions five- to seven-fold, and mesenchymal tumor risk was similar for *TERF2IP* and *POT1* despite a six-fold difference in telomere elongation. Shelterin proteins have functions beyond length control, including suppression of ATR and ATM signaling at telomeres and protection against telomere fusion.^3^ Partial loss of these functions may create chromosomal instability that adds to the proliferative advantage of long telomeres. Alternatively, leukocyte telomere length may understate elongation in target tissues. Either way, telomere length measurement alone will underestimate the risk carried by these variants.

The centrosome findings argue for caution. Neither core nor extended centrosome sets showed a directional pLoF signal for any outcome. Centrosome variants may confer risk specific to sarcoma or to early-onset disease, which an adult cohort recruited at ages 40 to 69 years cannot capture; until replicated, these genes should not be interpreted clinically.

There are clinical implications. Shelterin variants identified on sarcoma panels should prompt counseling on the established long-telomere spectrum (CLL and myeloma, thyroid cancer, melanoma),^27^ which should now include *TERF1*. The mesenchymal findings do not justify surveillance for lipoma or leiomyoma, which are common and benign, but they support considering shelterin genes as candidate predisposition genes in patients with leiomyosarcoma or liposarcoma, particularly with a personal or family history of the long-telomere spectrum.

This study has limitations. The AstraZeneca and Genebass analyses draw on overlapping UK Biobank participants; the second pipeline controls for analytic choices, not sampling, and independent replication is required. The portal export omitted results with *P* > .10, precluding a complete forest of null associations and formal heterogeneity testing across genes. Lipoma and leiomyoma were ascertained from hospital and self-report records and are underdiagnosed, which should bias toward the null. Several associations rest on fewer than ten carriers, particularly the TERF1 associations with MGUS, choroidal melanoma, and intra-abdominal lipoma, and nominal *P* values across many phenotypes carry a false-positive expectation, which we mitigated by prespecifying mesenchymal outcomes and requiring cross-gene concordance. Analyses were restricted to European ancestry. The observed-versus-predicted comparison assumes a log-linear MR effect that may not hold at the extreme elongation seen in carriers. Sarcoma ascertainment was inadequate for direct inference, as the null positive control shows.

In conclusion, germline shelterin loss of function predisposes to benign adipocytic and smooth muscle neoplasia at population scale, providing a lineage-specific rationale for its reported association with sarcoma. *TERF1* is a long-telomere cancer gene, and carrier risk exceeds what telomere length alone predicts for several cancers. Resolving sarcoma-specific penetrance will require sarcoma-enriched sequencing with histology-level phenotypes and tumor telomere maintenance status.

## Data Availability

All analyses used publicly available summary statistics; no individual-level data were accessed, and none are required to reproduce the results. Gene-level collapsing results came from the AstraZeneca PheWAS Portal (https://azphewas.com) and Genebass (https://genebass.org), both built on UK Biobank whole-exome sequencing, and outcome statistics for Mendelian randomization from FinnGen release 12.

Analysis code and the five summary-statistic extracts analyzed here are provided as Supplementary Material with this preprint, under the MIT License. A single command rebuilds Tables 1, 2, and 3 from those extracts and checks all 269 cells against the published values, including the permutation-calibrated gene-set statistics, which are deterministic under a fixed seed.

One boundary is worth stating. The Mendelian randomization estimates are distributed as results rather than recomputed by the repository: re-deriving them from instrument level requires the FinnGen outcome statistics for the telomere-length instruments, which are included, together with the published instrument effects of Codd et al.,^20^ which are not redistributed. The deposited file carries the full sensitivity suite, so the robustness claims above can be checked against it directly.

## Acknowledgments

This research used publicly released summary statistics from the UK Biobank Resource, analyzed through the AstraZeneca PheWAS Portal and Genebass, and from FinnGen. We thank the participants and investigators of those studies, and the teams that built and maintain the two portals.

## Author Contributions

Conceptualization: M.P.; Data Curation: M.P., B.D.L; Formal Analysis: M.P., B.D.L., K.H.C., Z.Z., L.Z., A.L., J.C.C.; Investigation: M.P., B.D.L., K.H.C., L.Z., A.L., J.C.C; Methodology: M.P., B.D.L., K.H.C., L.Z., A.L., J.C.C; Software: M.P., B.D.L., K.H.C., L.Z., A.L., J.C.C; Supervision: M.P.; Validation: M.P., B.D.L., K.H.C., L.Z., A.L., J.C.C; Writing-original draft: M.P.; Writing-review and editing: all authors.

## Ethics Declaration

This study analyzed only publicly available, de-identified summary statistics and did not involve human participants, identifiable data, or biological samples; institutional review board approval and informed consent were therefore not required for this analysis. The contributing studies obtained their own approvals and participant consent: UK Biobank has ethical approval from the North West Multi-centre Research Ethics Committee and all participants provided written informed consent, and FinnGen as described in its primary report. The research conformed to the principles of the Declaration of Helsinki.

## Declaration of AI and AI-assisted technologies in the writing process

During the preparation of this work the authors used a large language model (Claude; Anthropic, San Francisco, CA) to assist with retrieval of summary statistics, development of analysis code, statistical analysis, figure generation, and manuscript editing. The authors reviewed and verified the analyses, code, data sources, and cited references independently, edited the text, and take full responsibility for the content of the published article.

## Supplementary Material

**Supplementary Figure S1.**
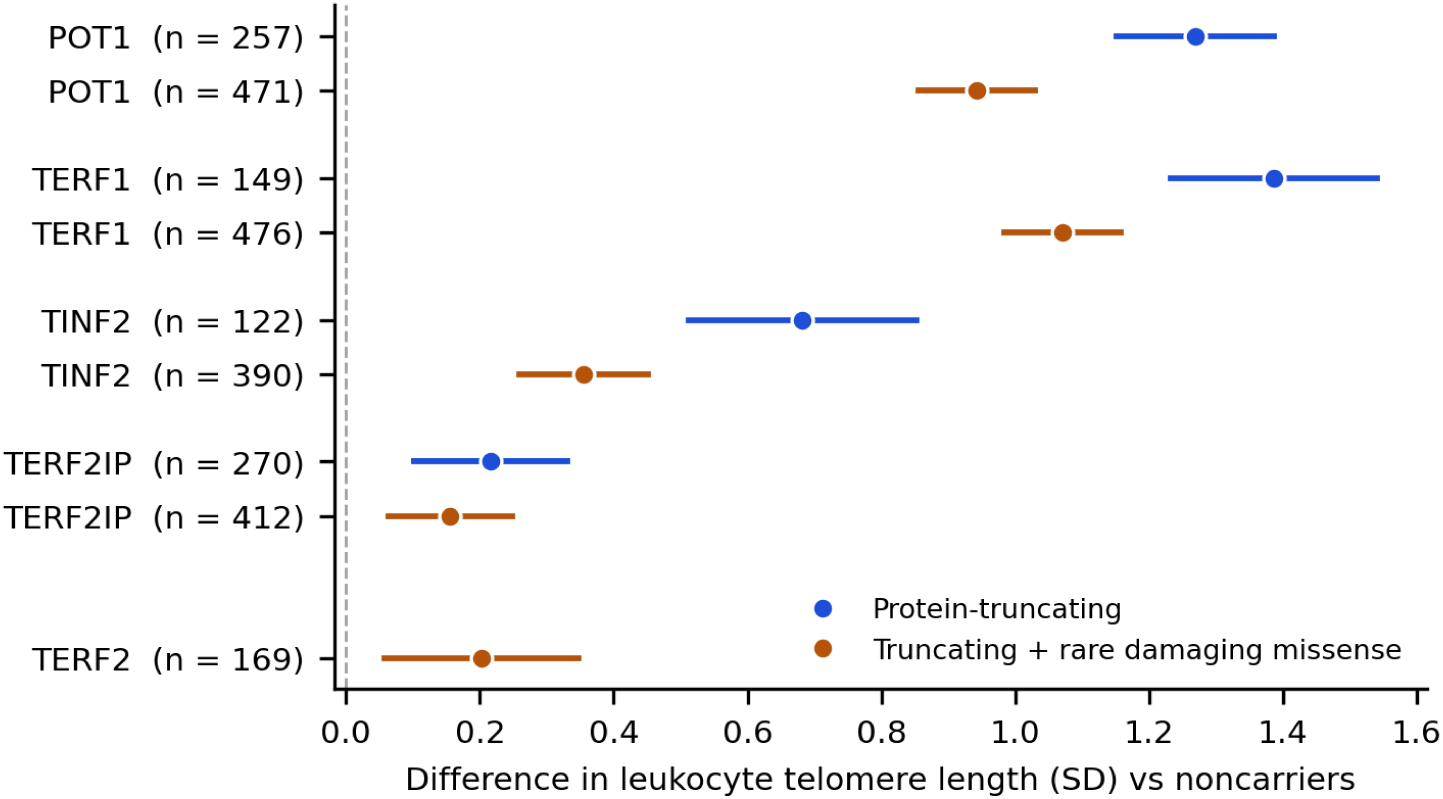
Rare shelterin variants lengthen leukocyte telomere length. Carrier minus noncarrier difference (SD) for PTV (blue) and PTV + RDM (orange); n, carriers.

**Supplementary Figure S2.**
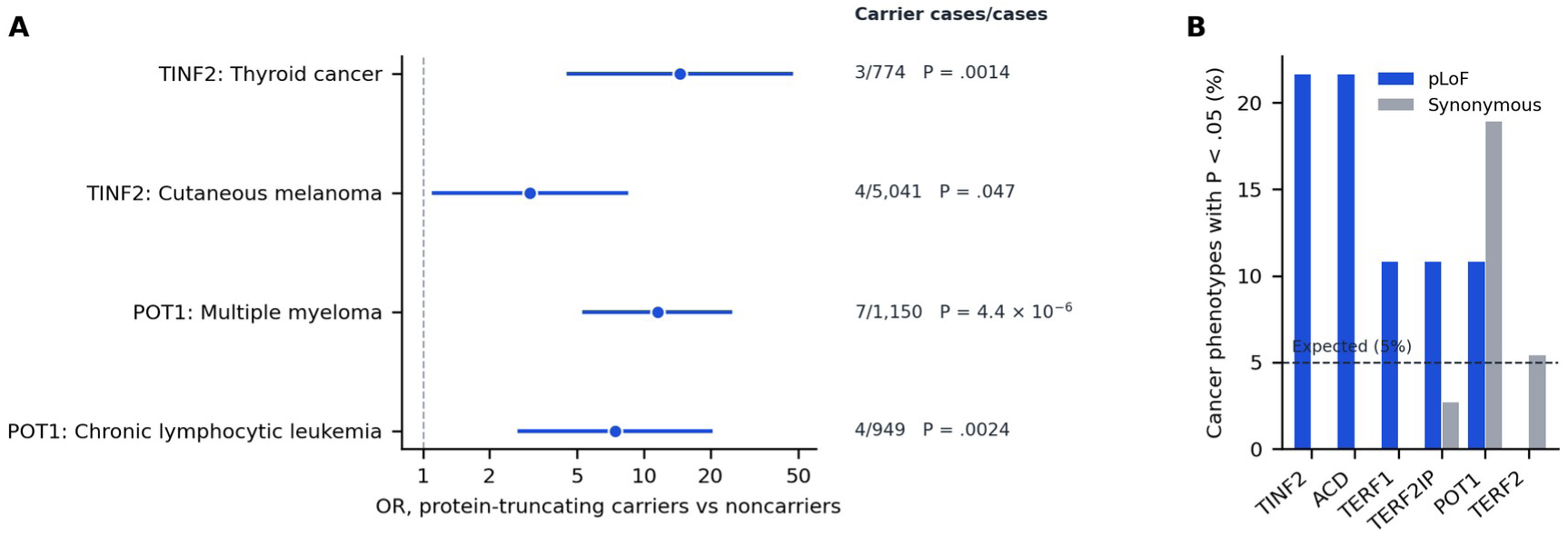
Reproduction of established *POT1* and *TINF2* cancer associations. (A) ORs for lead cancer registry associations. (B) Proportion of 37 ICD-10 cancer sites with SKAT-O *P* < .05 for pLoF (blue) and synonymous (gray) variants per shelterin gene (Genebass); dashed line, 5% null expectation.

**Supplementary Figure S3.**
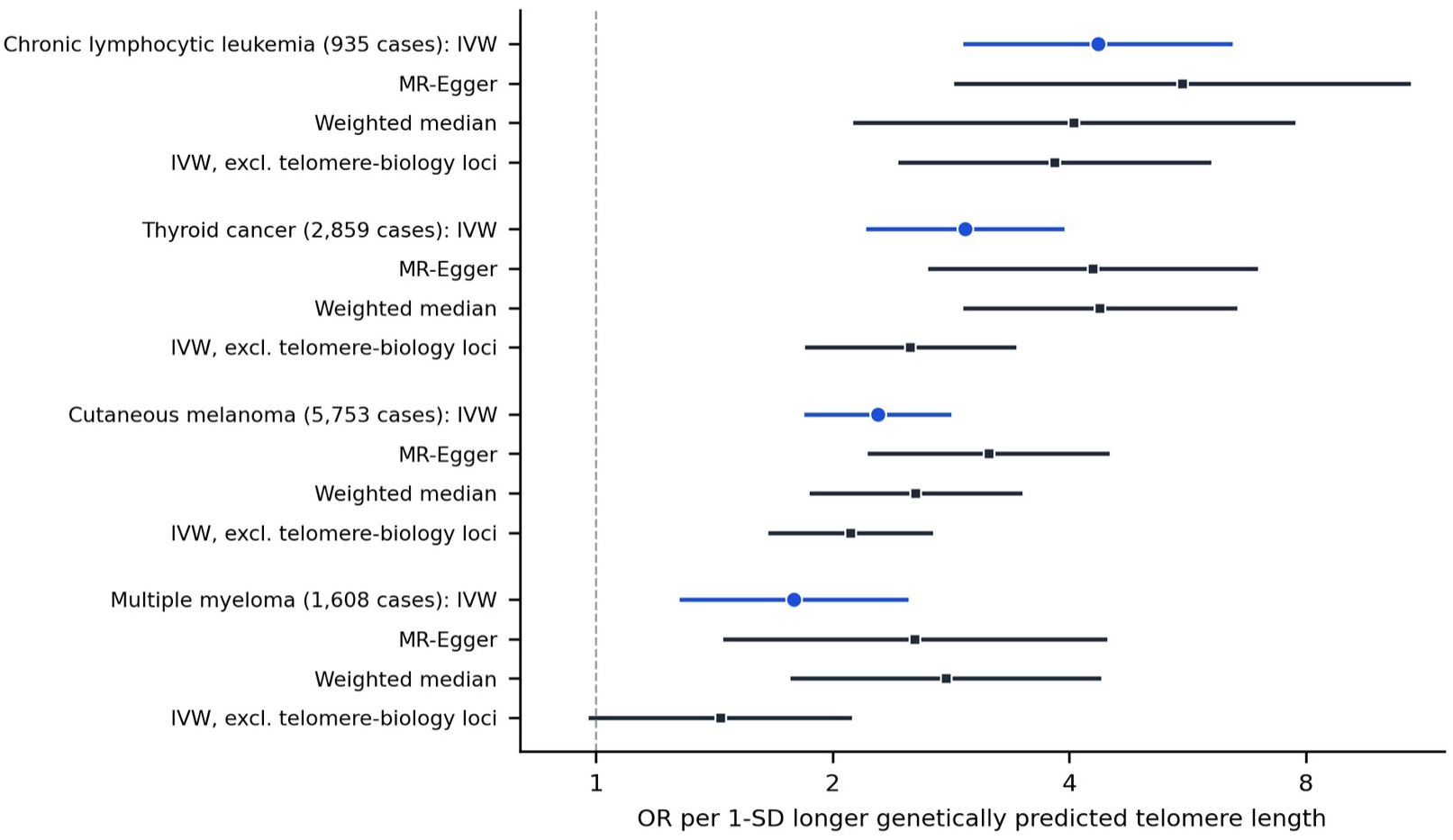
Mendelian randomization of leukocyte telomere length on chronic lymphocytic leukemia, thyroid cancer, cutaneous melanoma, and multiple myeloma (FinnGen R12): IVW (blue), MR-Egger, weighted median, and IVW excluding telomere-biology loci (black).

